# Diagnostic and prognostic value of hematological and immunological markers in COVID-19 infection: A meta-analysis of 6320 patients

**DOI:** 10.1101/2020.07.08.20141218

**Authors:** Rami M. Elshazli, Eman A Toraih, Abdelaziz Elgaml, Mohammed El-Mowafy, Mohamed El-Mesery, Mohamed Nasreldien Amin, Mohammad H Hussein, Mary T. Killackey, Manal S Fawzy, Emad Kandil

**Affiliations:** Department of Biochemistry and Molecular Genetics, Faculty of Physical Therapy, Horus University - Egypt, New Damietta, 34517, Egypt; Department of Surgery, Tulane University, School of Medicine, New Orleans, Louisiana, USA; Genetics Unit, Department of Histology and Cell Biology, Faculty of Medicine, Suez Canal University, Ismailia, Egypt; Department of Microbiology and Immunology, Faculty of Pharmacy, Mansoura University, Mansoura 35516, Egypt; Department of Microbiology, Faculty of Pharmacy, Horus University - Egypt, New Damietta, 34517, Egypt; Department of Biochemistry, Faculty of Pharmacy, Mansoura University, Mansoura 35516, Egypt; Tulane Transplant Institute, Tulane University, School of Medicine, New Orleans, LA, USA; Department of Medical Biochemistry, Faculty of Medicine, Suez Canal University, Ismailia, Egypt; Division of Endocrine and Oncologic Surgery, Department of Surgery, Tulane University, School of Medicine, New Orleans, LA, USA

**Keywords:** COVID-19, laboratory markers, mortality, prognosis, SARS-CoV-2

## Abstract

**Objective:** Evidence-based characterization of the diagnostic and prognostic value of the hematological and immunological markers related to the epidemic of Coronavirus Disease 2019 (COVID-19) is critical to understand the clinical course of the infection and to assess in development and validation of biomarkers.

**Methods:** Based on systematic search in Web of Science, PubMed, Scopus, and Science Direct up to April 22, 2020, a total of 52 eligible articles with 6,320 laboratory-confirmed COVID-19 cohorts were included. Pairwise comparison between severe *versus* mild disease, Intensive Care Unit (ICU) *versus* general ward admission, and expired *versus* survivors were performed for 36 laboratory parameters. The pooled standardized mean difference (SMD) and 95% confidence intervals (CI) were calculated using the DerSimonian Laird method/random effects model and converted to Odds ratio (OR). The decision tree algorithm was employed to identify the key risk factor(s) attributed to severe COVID-19 disease.

**Results:** Cohorts with elevated levels of white blood cells (WBCs) (OR=1.75), neutrophil count (OR=2.62), D-dimer (OR=3.97), prolonged prothrombin time (PT) (OR=1.82), fibrinogen (OR=3.14), erythrocyte sedimentation rate (OR=1.60), procalcitonin (OR=4.76), IL-6 (OR=2.10), and IL-10 (OR=4.93) had higher odds of progression to severe phenotype. Decision tree model (sensitivity=100%, specificity=81%) showed the high performance of neutrophil count at a cut-off value of more than 3.74□x10^9^/L for identifying patients at high risk of severe COVID□19. Likewise, ICU admission was associated with higher levels of WBCs (OR=5.21), neutrophils (OR=6.25), D-dimer (OR=4.19), and prolonged PT (OR=2.18). Patients with high IL-6 (OR=13.87), CRP (OR=7.09), D-dimer (OR=6.36), and neutrophils (OR=6.25) had the highest likelihood of mortality.

**Conclusions:** Several hematological and immunological markers, in particular neutrophilic count, could be helpful to be included within the routine panel for COVID-19 infection evaluation to ensure risk stratification and effective management.

## Introduction

Coronavirus disease – 2019 (COVID-19) is a disease that was detected in December 2019 in Wuhan, China, and led to the risk of mortality of about 2% [1]. This disease is caused due to infection with a recently arising zoonotic virus known as the Severe Acute Respiratory Syndrome Coronavirus 2 (SARS-CoV-2) [2]. Previously, infection with coronaviruses appeared in 2002 within China in the form of SARS-CoV, and it appeared later also in 2012 within Saudi Arabia that was known as Middle East Respiratory Syndrome (MERS-CoV) [3, 4]. All these coronaviruses are enveloped positive-strand RNA viruses that are isolated from bats that can be transferred from animals to humans, human to human, and animals to animals [5]. They share a similarity in the clinical symptoms in addition to specific differences that have been recently observed [5-7]. The symptoms of this disease appear with different degrees that start in the first seven days with mild symptoms such as fever, cough, shortness of breath, and fatigue [8]. Afterward, critical symptoms may develop in some patients involving dyspnea and pneumonia that require patient’s management in intensive care units to avoid the serious respiratory complications that may lead to death [9]. However, there are no specific symptoms to diagnose coronavirus infection, and accurate testing depends on the detection of the viral genome using the reverse transcription-polymerase chain reaction (RT-PCR) analysis [10].

Unfortunately, COVID-19 is not limited to its country of origin, but it has spread all over the world. Therefore, there is no wonder emerging research has been directed to provide information and clinical data of patients infected with this virus that may help to not only to the early detection in different patient categories, but it will also help in the characterization of the viral complications with other chronic diseases [1, 2, 6, 9]. However, there is no sufficient data that characterize the changes in the hematological and immunological parameters in COVID-19 patients. In the current comprehensive meta-analysis study, we aimed to analyze different hematological, inflammatory, and immunological markers in COVID-19 patients at different clinical stages in different countries that may help in the early detection of COVID-19 infection and to discriminate between severity status of the disease to decrease the death risk.

## Materials and Methods

### Search strategy

This current meta-analysis was carried out according to the Preferred Reporting Items for Systematic reviews and Meta-analysis (PRISMA) statement [11] (**Table S1**). Relevant literature was retrieved from Web of Science, PubMed, Scopus, and Science Direct search engines up to April 22, 2020. Our search strategy included the following terms: “Novel coronavirus 2019”, “2019 nCoV”, “COVID-19”, “Wuhan coronavirus,” “Wuhan pneumonia,” or “SARS-CoV-2”. Besides, we manually screened out the relevant potential article in the references selected. The above process was performed independently by three participants.

### Study selection

No time or language restriction was applied. Inclusion criteria were as follows: (1) Types of Studies: retrospective, prospective, observational, descriptive or case control studies reporting laboratory features of COVID-19 patients; (2) Subjects: diagnosed patients with COVID-19 (3) Exposure intervention: COVID-19 patients diagnosed with Real Time-Polymerase Chain Reaction, radiological imaging, or both; with hematological testing included: complete blood picture (white blood cells, neutrophil count, lymphocyte count, monocyte count, eosinophils count, basophils, red blood cells, hemoglobin, hematocrit, and platelet count), coagulation profile (prothrombin time, international normalized ratio, activated partial thromboplastin time, thrombin time, fibrinogen, and D-dimer) or immunological parameters including inflammatory markers (ferritin, erythrocyte sedimentation rate, procalcitonin, and C-reactive protein), immunoglobulins (IgA, IgG, and IgM), complement tests (C3 and C4), interleukins (IL-4, IL-6, IL-8, IL-10, IL-2R, and TNF-α), and immune cells (B lymphocytes, T lymphocytes, CD4^+^ T cells, and CD8^+^ T cells); and (4) Outcome indicator: the mean and standard deviation or median and interquartile range for each laboratory test. The following exclusion criteria were considered: (1) Case reports, reviews, editorial materials, conference abstracts, summaries of discussions, (2) Insufficient reported data information; or (3) *In vitro* or *in vivo* studies.

### Data abstraction

Four investigators separately conducted literature screening, data extraction, and literature quality evaluation, and any differences were resolved through another two reviewers. Information extracted from eligible articles in a predesigned form in excel, including the last name of the first author, date and year of publication, journal name, study design, country of the population, sample size, and quality assessment.

### Quality assessment

A modified version of the Newcastle-Ottawa scale (NOS) was adopted to evaluate the process in terms of queue selection, comparability of queues, and evaluation of results [12, 13]. The quality of the included studies was assessed independently by three reviewers, and disagreements were resolved by the process described above. Higher NOS scores showed a higher literature quality. NOS scores of at least six were considered high-quality literature.

### Statistical analysis

All data analysis was performed using OpenMeta[Analyst] [14] and comprehensive meta-analysis software version 3.0 [15]. First, a single-arm meta□analysis for laboratory tests was performed. The standardized mean difference (SMD) and 95%confidence intervals (CI) were used to estimate pooled results from studies. Medians and interquartile range were converted to mean and standard deviation (SD) using the following formulas: [Mean=(Q1+median+Q3)/3] and [SD=IQR/1.35], whereas, values reported in the articles as mean and 95%CI were estimated using the following formula [SD= √ N * (Upper limit of CI – Lower limit of CI)/3.92]. A continuous random-effect model was applied using the DerSimonian-Laird (inverse variance) method [16, 17].

Next, in the presence of individual patient data, single-armed observed values were converted to two-armed data to act as each other’s control group based on covariate information. Only studies investigating different outcomes were considered as potential matched pairs, and two-arm meta-analysis was applied to compare between mild *versus* severe COVID-19 infection (based on the results of the chest radiography, clinical examination, and symptoms), ICU admission *versus* general ward admission, and expired *versus* survivors. Meta-analysis for each outcome was processed using a random-effects model since heterogeneity among studies was expected. For severity pairwise comparison, estimates of SMD served as quantitative measures of the strength of evidence against the null hypothesis of no difference in the population between mild and severe COVID-19 manifestations. SMD of <0.2, 0.2-0.8, and >0.8 indicated mild, moderate, and severe strength. For ICU admission and survival analysis, overall effect size estimates in SMD were then converted to the odds ratio (OR) with 95%CI for better interpretation by clinical domains.

### Decision tree to identify predictors for poor outcomes

Using laboratory features for clinical prediction, the decision tree algorithm was employed to identify the key risk factors attributed to severe COVID-19 infection. The accuracy of the model was measured by the Area Under the Receiver Operating Characteristic (ROC) Curve (AUC), which depicts the true positive rate versus the false positive rate at various discrimination thresholds. The markers that have the highest AUC were identified, and the sensitivity and specificity of the cut-off threshold level were determined. R Studio was employed using the following packages: *tidyverse, magrittr, rpart, caret*, and *pROC*.

### Trial sequential analysis (TSA)

The statistical trustworthiness of this meta-analysis assessment was conducted using TSA through combining the cumulative sample sizes of all appropriate records with the threshold of statistical impact to diminish the accidental errors and enhance the intensity of expectations [18]. Two side trials with “type I error (α)” along with power set at 5% and 80% were employed. In the case of the “Z-curve” traverses the TSA monitoring boundaries, a reasonable degree of impact was accomplished, and no supplementary trials are crucial. Nevertheless, in case of the “Z-curve” failed to achieve the boundary limits, the estimated information size has not accomplished the required threshold to attract appropriate decisions and advance trials are mandatory. TSA platform (version 0.9.5.10 beta) was operated in the experiment.

### Assessment of heterogeneity and publication bias

After that, the heterogeneity was evaluated using Cochran’s Q statistic and quantified by using I^2^ statistics, which represents an estimation of the total variation across studies beyond chance. Articles were considered to have significant heterogeneity between studies when the p-value less than 0.1 or I^2^ greater than 50%. Subgroup analysis was performed based on the study sample size (≤50 patients compared to >50 patients) and the origin of patients (Wuhan city versus others). In addition, sensitivity analyses and meta-regression with the random-effects model using restricted maximum likelihood algorithm were conducted to explore potential sources of heterogeneity.

Finally, publication bias was assessed using a funnel plot and quantified using Begg’s and Mazumdar rank correlation with continuity correction and Egger’s linear regression tests. Asymmetry of the collected studies’ distribution by visual inspection or P-value < 0.1 indicated obvious publication bias [19]. The Duval and Tweedie’s trim and fill method’s assumption were considered to reduce the bias in pooled estimates [20].

## Results

### Literature search

A flowchart outlining the systematic review search results is described in **Fig 1A**. A total of 4752 records were identified through four major electronic databases till April 22, 2020 including Web of Science (n = 557), PubMed (n = 1688), Scopus (n = 1105) and Science Direct (n = 1402). Upon reviewing the retrieved articles, a total of 1230 records were excluded for duplication, and 3522 unique records were initially identified. Following screening of titles and abstracts, several studies were excluded for being case records (n = 44), review articles (n = 262), irrelevant publications (n = 1355), or editorial materials (n = 1809). The resulted 424 full-text publications were further assessed for eligibility, during which 372 records were removed for lacking sufficient laboratory data. Ultimately, a total of 52 eligible articles were included for the quantitative synthesis of this meta-analysis study, with 52 records represented single-arm analysis, 16 records represented two-arms severity analysis; meanwhile, 7 and 4 records were utilized for survival and ICU admission analyses, respectively.

**Fig 1.**
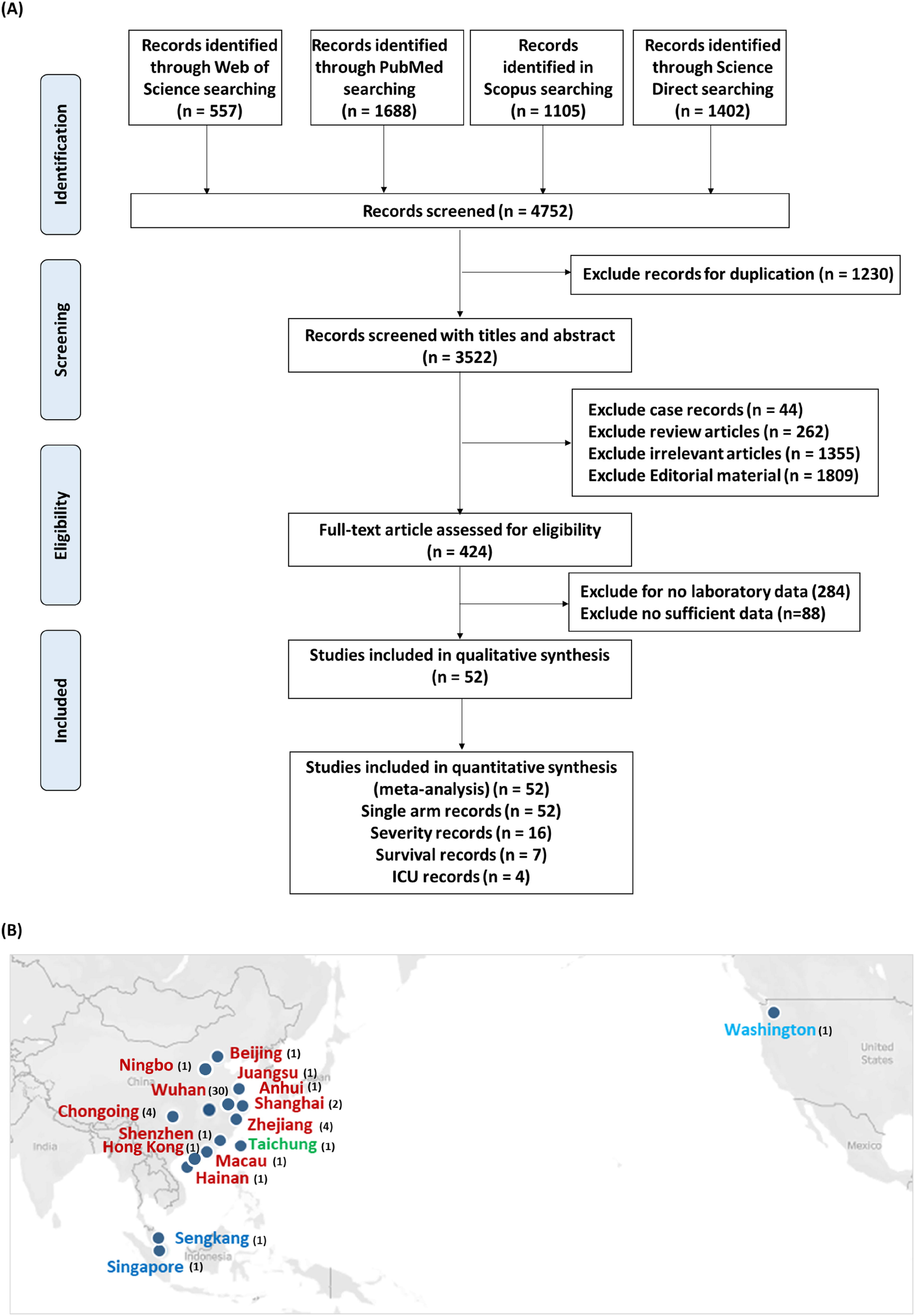
Literature search process. (A) Workflow for screening and selecting relevant articles. (B) Map showing the location of the studies. Studies conducted in China (red), Taiwan (green), Singapore (blue), and USA (light blue) are shown with the number of studies between brackets. Data source Tableau 2020.1 Desktop Professional Edition (https://www.tableau.com/).

### Characteristics of the included studies

Our review included 52 studies that were published from January 24 through April 22, 2020, including 48 articles from China [Wuhan (30), Chongqing (4), Zhejiang (4), Shanghai (2), Ningbo (1), Hong Kong (1), Shenzhen (1), Anhui (1), Macau (1), Hainan (1), Jiangsu (1), and Beijing (1)], two articles from Singapore [Singapore and Sengkang], one article from Taiwan [Taichung], and one article from USA [Washington] (**Fig 1B)**. The main characteristics of eligible studies are shown in **Table 1**. A total of 6320 patients with SARS□CoV□2 infection were enrolled across the articles. Most records (n = 47) were retrospective case studies, while other study design included two prospective cohort studies, one observational cohort study, one descriptive case series, and one case-control study. Our team stratified 36 different laboratory parameters into seven subclasses, including complete blood picture, coagulation profile, immunological markers, immunoglobulins, complement tests, interleukins, and immune cells, as previously described in the methodology. Regarding quality score assessment, 39 studies achieved a score higher than six out of a maximum of nine (high quality), while the remaining 13 studies earned a score equal or lower than six (low quality), as shown in **Table 1**.

**Table 1.**
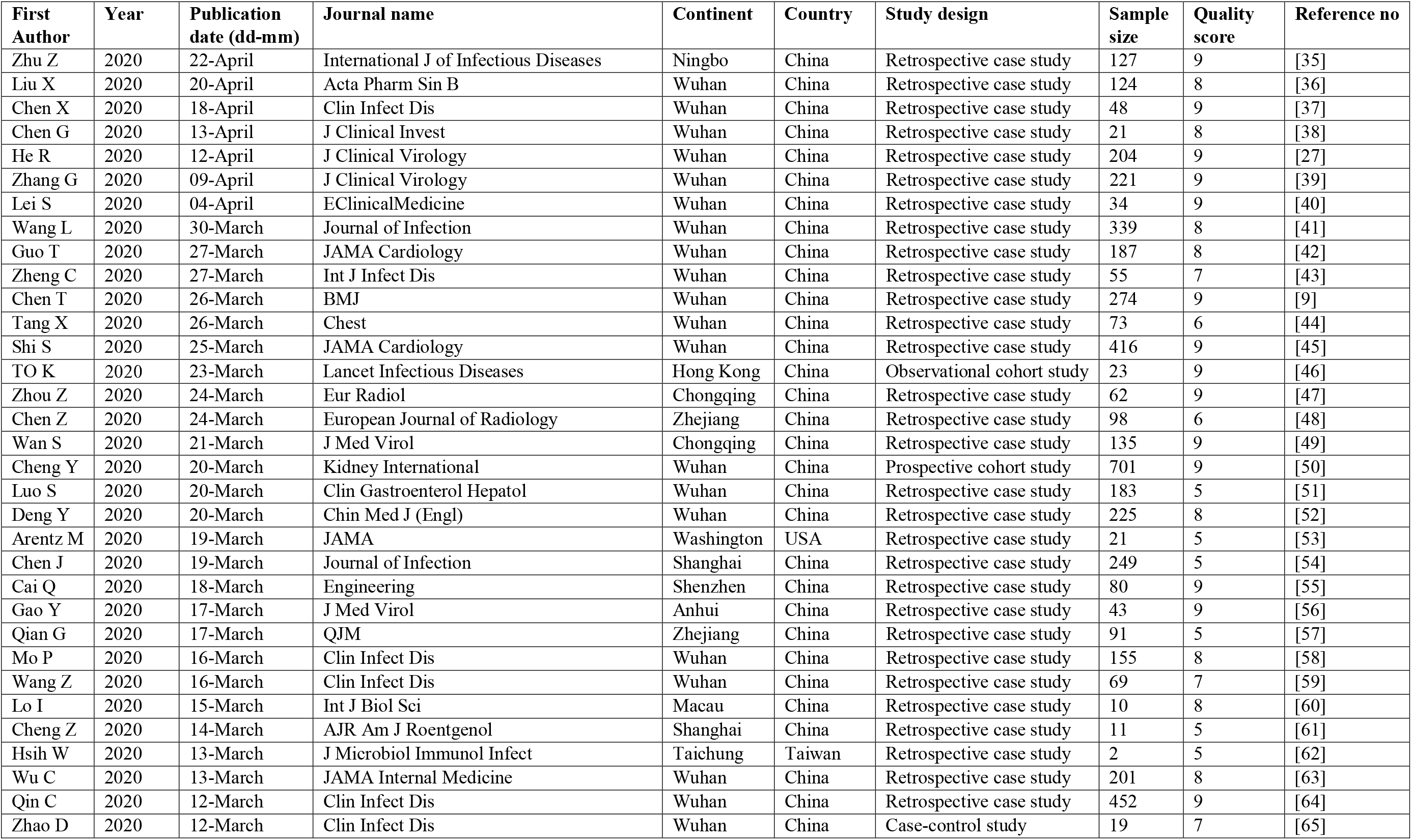
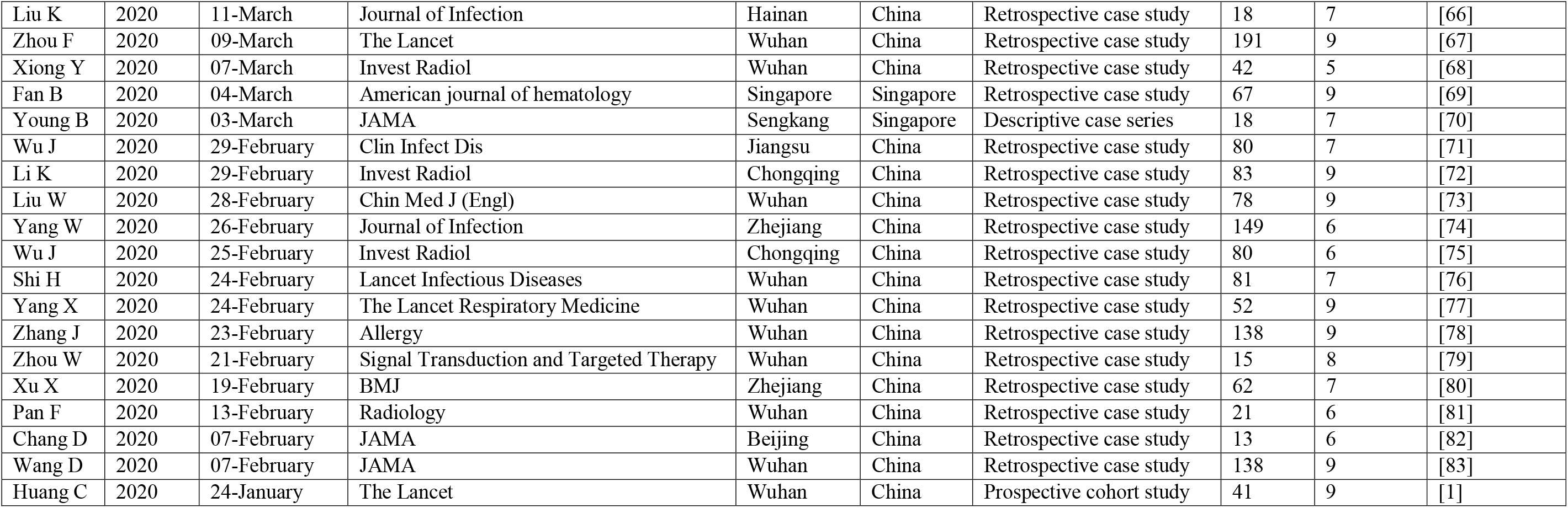
General characteristics of the included studies.

| First Author | Year | Publication date (dd-mm) | Journal name | Continent | Country | Study design | Sample size | Quality score | Reference no |
| --- | --- | --- | --- | --- | --- | --- | --- | --- | --- |
| Zhu Z | 2020 | 22-April | International J of Infectious Diseases | Ningbo | China | Retrospective case study | 127 | 9 | [35] |
| Liu X | 2020 | 20-April | Acta Pharm Sin B | Wuhan | China | Retrospective case study | 124 | 8 | [36] |
| Chen X | 2020 | 18-April | Clin Infect Dis | Wuhan | China | Retrospective case study | 48 | 9 | [37] |
| Chen G | 2020 | 13-April | J Clinical Invest | Wuhan | China | Retrospective case study | 21 | 8 | [38] |
| He R | 2020 | 12-April | J Clinical Virology | Wuhan | China | Retrospective case study | 204 | 9 | [27] |
| Zhang G | 2020 | 09-April | J Clinical Virology | Wuhan | China | Retrospective case study | 221 | 9 | [39] |
| Lei S | 2020 | 04-April | EClinicalMedicine | Wuhan | China | Retrospective case study | 34 | 9 | [40] |
| Wang L | 2020 | 30-March | Journal of Infection | Wuhan | China | Retrospective case study | 339 | 8 | [41] |
| Guo T | 2020 | 27-March | JAMA Cardiology | Wuhan | China | Retrospective case study | 187 | 8 | [42] |
| Zheng C | 2020 | 27-March | Int J Infect Dis | Wuhan | China | Retrospective case study | 55 | 7 | [43] |
| Chen T | 2020 | 26-March | BMJ | Wuhan | China | Retrospective case study | 274 | 9 | [9] |
| Tang X | 2020 | 26-March | Chest | Wuhan | China | Retrospective case study | 73 | 6 | [44] |
| Shi S | 2020 | 25-March | JAMA Cardiology | Wuhan | China | Retrospective case study | 416 | 9 | [45] |
| TO K | 2020 | 23-March | Lancet Infectious Diseases | Hong Kong | China | Observational cohort study | 23 | 9 | [46] |
| Zhou Z | 2020 | 24-March | Eur Radiol | Chongqing | China | Retrospective case study | 62 | 9 | [47] |
| Chen Z | 2020 | 24-March | European Journal of Radiology | Zhejiang | China | Retrospective case study | 98 | 6 | [48] |
| Wan S | 2020 | 21-March | J Med Virol | Chongqing | China | Retrospective case study | 135 | 9 | [49] |
| Cheng Y | 2020 | 20-March | Kidney International | Wuhan | China | Prospective cohort study | 701 | 9 | [50] |
| Luo S | 2020 | 20-March | Clin Gastroenterol Hepatol | Wuhan | China | Retrospective case study | 183 | 5 | [51] |
| Deng Y | 2020 | 20-March | Chin Med J (Engl) | Wuhan | China | Retrospective case study | 225 | 8 | [52] |
| Arentz M | 2020 | 19-March | JAMA | Washington | USA | Retrospective case study | 21 | 5 | [53] |
| Chen J | 2020 | 19-March | Journal of Infection | Shanghai | China | Retrospective case study | 249 | 5 | [54] |
| Cai Q | 2020 | 18-March | Engineering | Shenzhen | China | Retrospective case study | 80 | 9 | [55] |
| Gao Y | 2020 | 17-March | J Med Virol | Anhui | China | Retrospective case study | 43 | 9 | [56] |
| Qian G | 2020 | 17-March | QJM | Zhejiang | China | Retrospective case study | 91 | 5 | [57] |
| Mo P | 2020 | 16-March | Clin Infect Dis | Wuhan | China | Retrospective case study | 155 | 8 | [58] |
| Wang Z | 2020 | 16-March | Clin Infect Dis | Wuhan | China | Retrospective case study | 69 | 7 | [59] |
| Lo I | 2020 | 15-March | Int J Biol Sci | Macau | China | Retrospective case study | 10 | 8 | [60] |
| Cheng Z | 2020 | 14-March | AJR Am J Roentgenol | Shanghai | China | Retrospective case study | 11 | 5 | [61] |
| Hsieh W | 2020 | 13-March | J Microbiol Immunol Infect | Taichung | Taiwan | Retrospective case study | 2 | 5 | [62] |
| Wu C | 2020 | 13-March | JAMA Internal Medicine | Wuhan | China | Retrospective case study | 201 | 8 | [63] |
| Qin C | 2020 | 12-March | Clin Infect Dis | Wuhan | China | Retrospective case study | 452 | 9 | [64] |
| Zhao D | 2020 | 12-March | Clin Infect Dis | Wuhan | China | Case-control study | 19 | 7 | [65] |
| Liu K | 2020 | 11-March | Journal of Infection | Hainan | China | Retrospective case study | 18 | 7 | [66] |
| Zhou F | 2020 | 09-March | The Lancet | Wuhan | China | Retrospective case study | 191 | 9 | [67] |
| Xiong Y | 2020 | 07-March | Invest Radiol | Wuhan | China | Retrospective case study | 42 | 5 | [68] |
| Fan B | 2020 | 04-March | American journal of hematology | Singapore | Singapore | Retrospective case study | 67 | 9 | [69] |
| Young B | 2020 | 03-March | JAMA | Sengkang | Singapore | Descriptive case series | 18 | 7 | [70] |
| Wu J | 2020 | 29-February | Clin Infect Dis | Jiangsu | China | Retrospective case study | 80 | 7 | [71] |
| Li K | 2020 | 29-February | Invest Radiol | Chongqing | China | Retrospective case study | 83 | 9 | [72] |
| Liu W | 2020 | 28-February | Chin Med J (Engl) | Wuhan | China | Retrospective case study | 78 | 9 | [73] |
| Yang W | 2020 | 26-February | Journal of Infection | Zhejiang | China | Retrospective case study | 149 | 6 | [74] |
| Wu J | 2020 | 25-February | Invest Radiol | Chongqing | China | Retrospective case study | 80 | 6 | [75] |
| Shi H | 2020 | 24-February | Lancet Infectious Diseases | Wuhan | China | Retrospective case study | 81 | 7 | [76] |
| Yang X | 2020 | 24-February | The Lancet Respiratory Medicine | Wuhan | China | Retrospective case study | 52 | 9 | [77] |
| Zhang J | 2020 | 23-February | Allergy | Wuhan | China | Retrospective case study | 138 | 9 | [78] |
| Zhou W | 2020 | 21-February | Signal Transduction and Targeted Therapy | Wuhan | China | Retrospective case study | 15 | 8 | [79] |
| Xu X | 2020 | 19-February | BMJ | Zhejiang | China | Retrospective case study | 62 | 7 | [80] |
| Pan F | 2020 | 13-February | Radiology | Wuhan | China | Retrospective case study | 21 | 6 | [81] |
| Chang D | 2020 | 07-February | JAMA | Beijing | China | Retrospective case study | 13 | 6 | [82] |
| Wang D | 2020 | 07-February | JAMA | Wuhan | China | Retrospective case study | 138 | 9 | [83] |
| Huang C | 2020 | 24-January | The Lancet | Wuhan | China | Prospective cohort study | 41 | 9 | [1] |

### Pooled estimates of laboratory parameters: Single-arm Meta-analysis

The final pooled estimates of single-arm meta-analysis included 52 eligible articles. The pooled mean of laboratory parameters and 95%CI among SARS-CoV-2 infected patients, including hematological, immunological, and inflammatory variables, is illustrated in **Table 2**. Our results depicted a wide variability between studies for each laboratory marker. Apart from immunoglobulins, IL-2R, and IL-8, significant heterogeneity was observed. Subgroup analysis by sample size and city of origin and sensitivity analysis failed to reveal the source of variation for each parameter. Additionally, meta-regression also rendered insignificant results.

**Table 2.** **Pooled estimates of single-arm meta-analysis for laboratory parameters in COVID-19 patients**

| Laboratory testing | Number studies | Sample size | Estimate | 95% CI | P-value | Q | P-value | I<sup>2</sup> | T<sup>2</sup> |
| --- | --- | --- | --- | --- | --- | --- | --- | --- | --- |
| **CBC** |  |  |  |  |  |  |  |  |  |
| White blood cells | 47 | 5967 | 5.82 | 5.24, 6.40 | <0.001 | 7136.1 | <0.001 | 99.35 | 3.83 |
| Neutrophil count | 31 | 3814 | 3.70 | 3.48, 3.92 | <0.001 | 525.8 | <0.001 | 93.9 | 0.31 |
| Lymphocyte count | 45 | 6017 | 0.99 | 0.91, 1.08 | <0.001 | 7645.2 | <0.001 | 99.3 | 0.07 |
| Monocyte count | 18 | 2586 | 0.42 | 0.39, 0.44 | <0.001 | 263.7 | <0.001 | 93.5 | 0.003 |
| Eosinophils count | 4 | 546 | 0.02 | 0.01, 0.024 | <0.001 | 10.6 | 0.014 | 71.6 | 0.0 |
| Red blood cells | 2 | 507 | 4.42 | 3.81, 4.67 | <0.001 | 50.8 | <0.001 | 98.03 | 0.095 |
| Hemoglobin | 26 | 3114 | 129.1 | 125.0, 133.3 | <0.001 | 1504.3 | <0.001 | 98.3 | 103.4 |
| Platelet count | 34 | 4347 | 178.4 | 171.9, 184.9 | <0.001 | 390.2 | <0.001 | 91.5 | 273.5 |
| **Coagulation profile** |  |  |  |  |  |  |  |  |  |
| Prothrombin time | 22 | 3287 | 12.38 | 11.8, 12.9 | <0.001 | 3415.7 | <0.001 | 99.3 | 1.905 |
| APTT | 19 | 3023 | 31.8 | 30.2, 33.4 | <0.001 | 1312.1 | <0.001 | 98.6 | 11.96 |
| Thrombin time | 2 | 754 | 21.9 | 8.29, 35.57 | 0.002 | 1908.1 | <0.001 | 99.94 | 96.86 |
| D-dimer | 27 | 3857 | 1.25 | 0.67, 1.82 | <0.001 | 40947.5 | <0.001 | 99.9 | 2.22 |
| Fibrinogen | 2 | 781 | 2.45 | 0.61, 4.29 | 0.009 | 46.19 | <0.001 | 97.83 | 1.729 |
| **Inflammatory markers** |  |  |  |  |  |  |  |  |  |
| Ferritin | 8 | 528 | 889.5 | 773.2, 1005.7 | <0.001 | 16.61 | 0.020 | 57.8 | 14138.9 |
| ESR | 13 | 1013 | 37.85 | 29.07, 46.6 | <0.001 | 692.4 | <0.001 | 98.26 | 239.7 |
| Procalcitonin | 25 | 3010 | 0.10 | 0.07, 0.12 | <0.001 | 3913.6 | <0.001 | 99.3 | 0.003 |
| C-reactive protein | 36 | 4409 | 28.11 | 24.7, 31.4 | <0.001 | 3432.1 | <0.001 | 98.9 | 79.35 |
| **Immunoglobulins** |  |  |  |  |  |  |  |  |  |
| IgA | 2 | 101 | 2.21 | 2.15, 2.27 | <0.001 | 0.089 | 0.76 | 0.0 | 0.0 |
| IgG | 2 | 101 | 11.54 | 11.2, 11.8 | <0.001 | 1.88 | 0.17 | 46.9 | 0.023 |
| IgM | 2 | 101 | 1.00 | 0.96, 1.04 | <0.001 | 1.11 | 0.29 | 10.32 | 0.0 |
| **Complement test** |  |  |  |  |  |  |  |  |  |
| C3 | 2 | 101 | 0.95 | 0.80, 1.10 | <0.001 | 28.02 | <0.001 | 96.43 | 0.011 |
| C4 | 2 | 101 | 0.24 | 0.21, 0.27 | <0.001 | 28.08 | <0.001 | 96.44 | 0.0 |
| **Interleukins** |  |  |  |  |  |  |  |  |  |
| IL-2R | 2 | 101 | 762.3 | 732.4, 792.2 | <0.001 | 0.33 | 0.56 | 0.0 | 0.0 |
| IL-4 | 2 | 276 | 2.98 | 1.09, 4.87 | 0.002 | 958.765 | <0.001 | 99.9 | 1.85 |
| IL-6 | 12 | 926 | 11.56 | 9.82, 13.3 | <0.001 | 144.7 | <0.001 | 92.4 | 6.19 |
| IL-8 | 2 | 101 | 18.4 | 17.08, 19.84 | <0.001 | 1.54 | 0.21 | 35.3 | 0.39 |
| IL-10 | 3 | 292 | 6.33 | 4.39, 8.27 | <0.001 | 133.1 | <0.001 | 98.4 | 2.89 |
| TNF-α | 3 | 292 | 6.72 | 1.33, 12.12 | 0.015 | 2933.6 | <0.001 | 99.9 | 22.7 |
| **Immune cells** |  |  |  |  |  |  |  |  |  |
| CD4<sup>+</sup> T cells | 6 | 296 | 361.1 | 254.0, 468.2 | <0.001 | 88.7 | <0.001 | 94.3 | 15973.1 |
| CD8<sup>+</sup> T cells | 5 | 285 | 219.6 | 157.1, 282.0 | <0.001 | 46.17 | <0.001 | 91.3 | 4437.2 |
| T lymphocytes | 2 | 167 | 704.3 | 254.5, 1154.0 | 0.002 | 27.6 | <0.001 | 96.3 | 101500 |

Test of association: standardized mean difference, Random model. 95% CI: 95% confidence interval, Q statistic: a measure of weighted squared deviations that denotes the ratio of the observed variation to the within-study error, $I^2$ : the ratio of true heterogeneity to total observed variation, $T^2$ : Tau squared, and it is referred to the extent of variation among the effects observed in different studies. Laboratory markers (INR and B lymphocytes) were reported in only one study thus were not shown. CBC: Complete blood picture, APTT: Activated partial thromboplastin time, ESR: Erythrocyte sedimentation rate. Ig: immunoglobulin, IL-2R: Interleukin-2 receptor, TNF-α : tumor necrosis factor-alpha.

### Pooled estimates of laboratory parameters according to disease severity: Pairwise Meta-analysis

Two-arms meta-analyses were then conducted for three pairwise comparisons; (1) Severe *versus* mild COVID, (2) ICU admitted patients *versus* the general ward, and (3) Expired *versus* survivors (**Table 3)**.

**Table 3.**
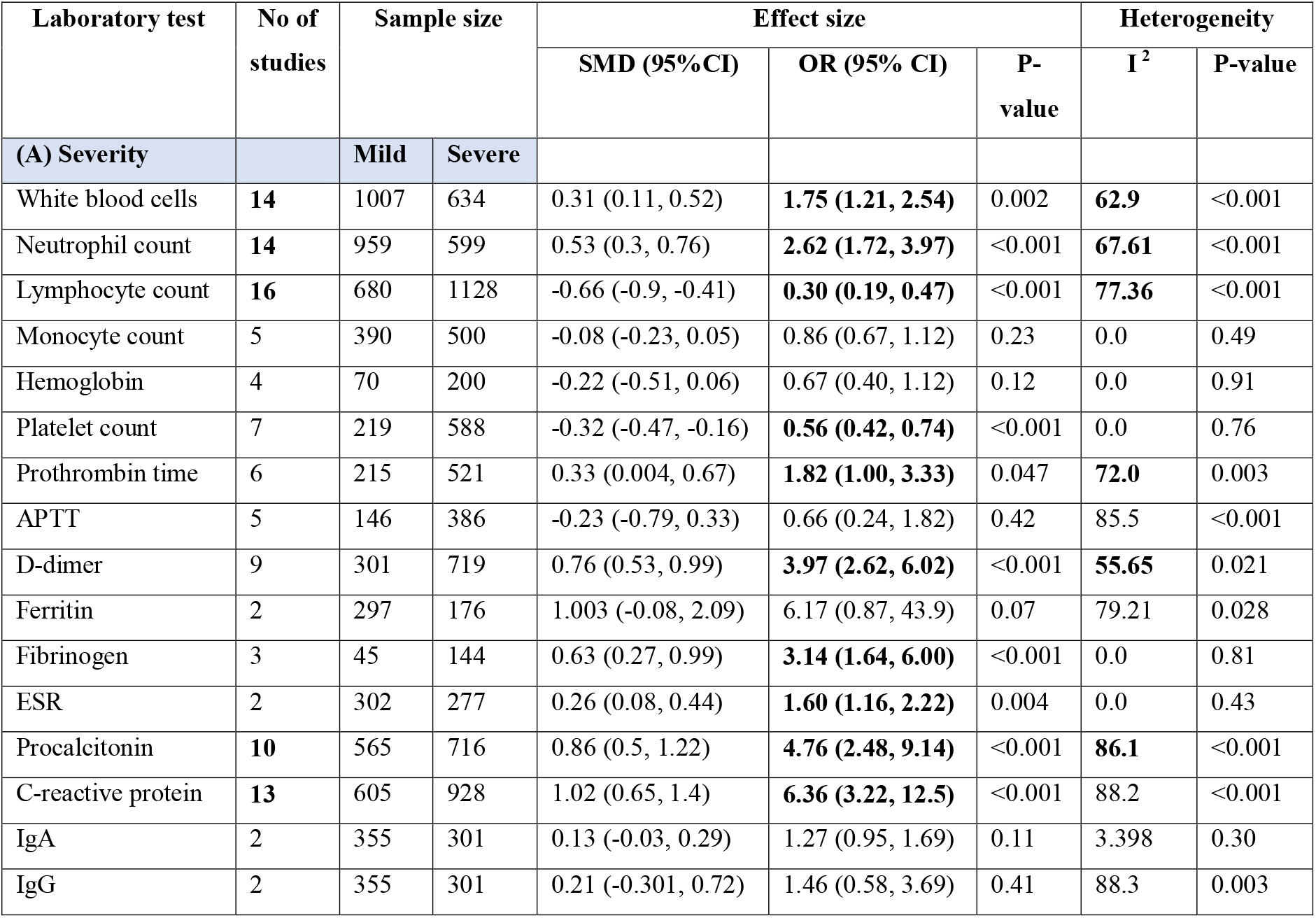
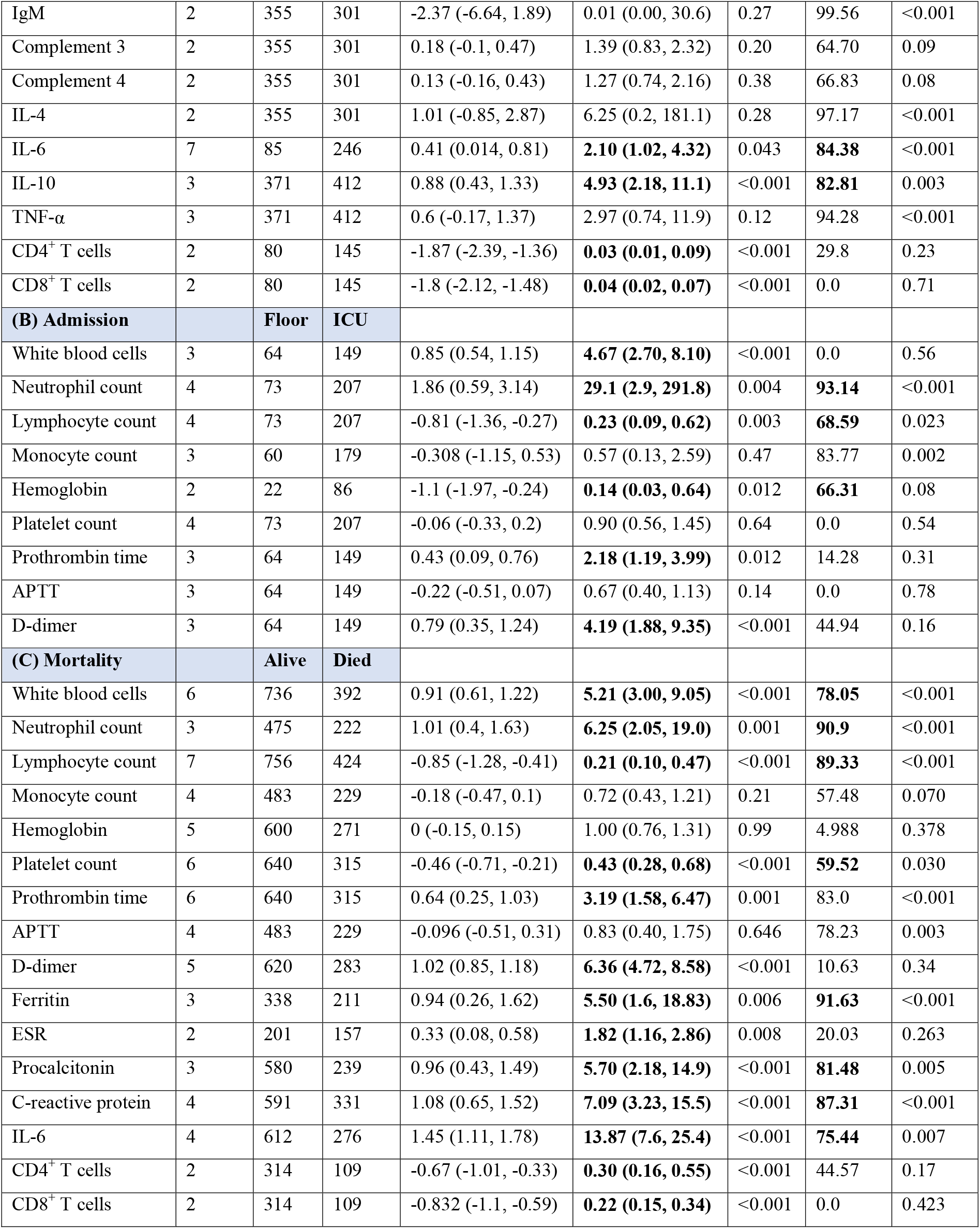
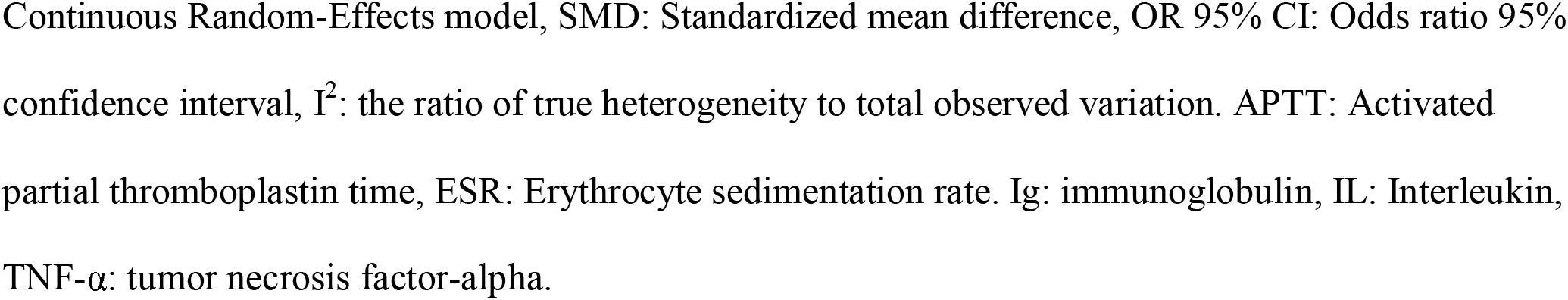
**Pooled estimates of two-arms meta-analysis for laboratory parameters in COVID-19 patients**.

Laboratory parameters of 16 eligible records were utilized to compare between severe and non-severe patients. Severe cohorts were more likely to have high blood levels of white blood cells (OR = 1.75, 95%CI = 1.21 - 2.54, *p* = 0.002), neutrophil count (OR = 2.62, 95%CI = 1.72 - 3.97, *p* <0.001), prothrombin time (OR = 1.82, 95%CI = 1.00 - 3.33, *p* = 0.047), D-dimer (OR = 3.97, 95%CI = 2.62 - 6.02, *p* <0.001), fibrinogen (OR = 3.14, 95%CI = 1.64 - 6.00, *p* <0.001), erythrocyte sedimentation rate (OR = 1.60, 95%CI = 1.16 - 2.22, *p* <0.001), procalcitonin (OR = 4.76, 95%CI = 2.48 - 9.14, *p* <0.001), IL-6 (OR = 2.10, 95%CI = 1.02 - 4.32, *p* = 0.043), and IL-10 (OR = 4.93, 95%CI = 2.18 - 11.1, *p* <0.001). In contrast, patients with normal lymphocyte count (OR = 0.30, 95%CI = 0.19 - 0.47, *p* <0.001), platelet count (OR = 0.56, 95%CI = 0.42 - 0.74, *p* <0.001), CD4^+^ T cells (OR = 0.04, 95%CI = 0.02 - 0.07, *p* <0.001), and CD8^+^ T cells (OR = 0.03, 95%CI = 0.01 - 0.09, *p* <0.001) were less likely to develop severe form of COVID-19 disease (**Table 3A)**.

Significant heterogeneity was observed in eight of these parameters, namely WBC (I^2^ = 62.9%, *p* <0.001), neutrophil count (I^2^ = 67.6%, *p* <0.001), lymphocyte count (I^2^ = 77.4%, *p* <0.001), prothrombin time (I^2^ = 72%, *p* = 0.003), D-dimers (I^2^ = 55.6%, *p* = 0.021), procalcitonin (I^2^ = 86.1%, *p* <0.001), IL-6 (I^2^ = 84.4%, *p* <0.001), and IL-10 (I^2^ = 82.8%, *p* = 0.003).

### Pooled estimates of laboratory parameters according to ICU admission: Pairwise Meta-analysis

A total of 4 eligible articles were recognized to include laboratory features of ICU and floor patients. Our data revealed having elevated levels of WBCs (OR = 5.21, 95%CI = 3.0 – 9.05, *p* <0.001), neutrophils (OR = 6.25, 95%CI = 2.05 – 19.0, *p* = 0.001), D-dimer (OR = 4.19, 95%CI = 1.88 - 9.35, *p* <0.001), and prolonged prothrombin time (OR = 2.18, 95%CI = 1.19 - 3.99, *p* =0.012) were associated with increased odds of ICU admission, while normal lymphocyte count (OR = 0.23, 95%CI = 0.09 - 0.62, *p* = 0.003) and hemoglobin (OR = 0.14, 95%CI = 0.03 - 0.64, *p* = 0.012) conferred lower risk of ICU admission (**Table 3B)**.

Remarkable heterogeneity was obvious in studies of neutrophil count (I^2^ = 93.1%, *p* <0.001), lymphocyte count (I^2^ = 68.5%, *p* = 0.023), and hemoglobin (I^2^ = 66.3%, *p* = 0.08). These parameters were enclosed in two to four studies; therefore, further tracing for the source of heterogeneity was not applicable.

### Pooled estimates of laboratory parameters according to mortality: Pairwise Meta-analysis

Of the included articles, 7 studies contained separate results for laboratory testing in survival *versus* expired patients. As depicted in **Table 3C**, our data revealed increased odds of having elevated levels of WBC (OR = 5.21, 95%CI = 3.0 – 9.05, *p* <0.001), neutrophils (OR = 6.25, 95%CI = 2.05 – 19.0, *p* = 0.001), prothrombin time (OR = 3.19, 95%CI = 1.58 – 6.47, *p* = 0.001), D-dimer (OR = 6.36, 95%CI = 4.72 - 8.58, *p* <0.001), ferritin (OR = 5.50, 95%CI = 1.6 - 18.8, *p* = 0.006), ESR (OR = 1.82, 95%CI = 1.16 - 2.86, *p* = 0.008), procalcitonin (OR = 5.70, 95%CI = 2.18 - 14.9, *p* <0.001), CRP (OR = 7.09, 95%CI = 3.23 - 15.5, *p* <0.001), and IL-6 (OR = 13.87, 95%CI = 7.6 - 25.4, *p* <0.001) in expired cases. However, patients with normal lymphocyte count (0.21 (0.10, 0.47, p <0.001), platelet count (0.43 (0.28, 0.68, p <0.001), CD4^+^ T cells (OR = 0.30 (0.16, 0.55, p <0.001), and CD8^+^ T cells (OR = 0.22 (0.15, 0.34, p <0.001) had higher chance of survival (**Table 3C)**.

Considerable heterogeneity was also noted in some of these parameters, namely WBC (I^2^ = 78.0%, *p* <0.001), neutrophilic count (I^2^ = 90.9%, *p* <0.001), lymphocyte count (I^2^ = 89.3%, *p* <0.001), platelet count (I^2^ = 59.5%, *p* = 0.030), ferritin (I^2^ = 91.6%, *p* <0.001), procalcitonin (I^2^ = 81.5%, *p* = 0.005), CRP (I^2^ = 87.3%, *p* <0.001), and IL-6 (I^2^ = 75.4%, *p* = 0.007). Given the small number of enrolled studies with discriminated data on patients who survived or died, we failed to identify the source of heterogeneity.

### Subgroup and sensitivity analysis

For the studies which included a comparison between mild and severe patients, subgroup and sensitivity analyses were performed for five laboratory markers (WBC, neutrophil count, lymphocyte count, procalcitonin, and CRP). First, to identify how each study affects the overall estimate of the rest of the studies, we performed leave-one-out sensitivity analyses. Results did not contribute to give explanations to heterogeneity. In contrast, subgroup analysis revealed homogeneity with certain categorizations. For WBCs lab results, heterogeneity was resolved on stratification by the origin of study population [Wuhan population: I^2^ = 73.4%, *p* = 0.002, other cities: I^2^ = 0%, *p* = 0.53] and month of publication [April: I^2^ = 74.5%, *p* = 0.001, February/March: I^2^ = 47.5%, *p* = 0.06]. Regarding neutrophilic count, the variance in the results resolved in articles with large sample size >50 patients (I^2^= 46.2%, *p* = 0.06). Moreover, the degree of dissimilarities of procalcitonin results found in different studies was ameliorated in April publications (I^2^ = 41.5%, *p* = 0.16) and in those with low sample size (I^2^ = 0%, *p* = 0.80). Similarly, homogeneity was generated in CRP results in articles with low sample size (I^2^ = 0%, *p* = 0.58) (**Table 4)**.

**Table 4.** **Tracing the source of heterogeneity of laboratory markers in studies comparing mild and severe COVID-19 patients**

| Lab test | Feature | Categories | Count of studies | Pooled estimates |  |  |  | Heterogeneity |  | Meta-regression |  |  |  |
| --- | --- | --- | --- | --- | --- | --- | --- | --- | --- | --- | --- | --- | --- |
|  |  |  |  | SMD | LL | UL | P-value | I<sup>2</sup> | P-value | Coefficient | LL | UL | P-value |
| **White blood cells** | Overall |  | 14 | 0.317 | 0.113 | 0.52 | 0.002 | 62.90% | 0.001 |  |  |  |  |
|  | Origin of patients | Others | 8 | 0.113 | -0.083 | 0.308 | 0.26 | 0% | 0.53 | Reference |  |  |  |
|  |  | Wuhan | 6 | 0.490 | 0.198 | 0.783 | 0.00 | 73.40% | 0.002 | 0.31 | 0.03 | 0.58 | 0.029 |
|  | Sample size | ≤50 | 5 | 0.164 | -0.553 | 0.881 | 0.65 | 71.30% | 0.007 | Reference |  |  |  |
|  |  | >50 | 9 | 0.387 | 0.208 | 0.566 | <0.001 | 52.60% | 0.031 | 0.30 | -0.10 | 0.72 | 0.14 |
|  | Publication month | Feb/Mar | 8 | 0.251 | 0.039 | 0.464 | 0.021 | 47.50% | 0.06 | Reference |  |  |  |
|  |  | April | 6 | 0.445 | 0.005 | 0.884 | 0.047 | 74.50% | 0.001 | 0.11 | -0.16 | 0.38 | 0.43 |
| **Neutrophils** | Overall |  | 14 | 0.534 | 0.306 | 0.762 | <0.001 | 67.62% | <0.001 |  |  |  |  |
|  | Origin of patients | Others | 8 | 0.439 | 0.139 | 0.740 | 0.004 | 50.88% | 0.047 | Reference |  |  |  |
|  |  | Wuhan | 6 | 0.632 | 0.280 | 0.985 | <0.001 | 78.29% | <0.001 | 0.045 | -0.21 | 0.30 | 0.20 |
|  | Sample size | ≤50 | 5 | 0.286 | -0.503 | 1.076 | 0.47 | 75.94% | 0.002 | Reference |  |  |  |
|  |  | >50 | 9 | 0.65 | 0.472 | 0.828 | <0.001 | 46.2% | 0.06 | 0.606 | 0.20 | 1.01 | 0.003 |
|  | Publication month | Feb/Mar | 8 | 0.428 | 0.181 | 0.675 | <0.001 | 54.4% | 0.032 | Reference |  |  |  |
|  |  | April | 6 | 0.709 | 0.273 | 1.44 | 0.001 | 73.19% | 0.002 | 0.312 | 0.06 | 0.55 | 0.014 |
| **Lymphocytes** | Overall |  | 16 | -0.663 | -0.909 | -0.417 | <0.001 | 77.36% | <0.001 |  |  |  |  |
|  | Origin of patients | Others | 9 | -0.626 | -0.962 | -0.291 | <0.001 | 66.51% | 0.002 | Reference |  |  |  |
|  |  | Wuhan | 7 | -0.710 | 1.097 | -0.323 | <0.001 | 85.72% | <0.001 | 0.092 | -0.31 | 0.49 | 0.64 |
|  | Sample size | ≤50 | 5 | -0.506 | -1.169 | 0.156 | 0.13 | 66.1% | 0.019 | Reference |  |  |  |
|  |  | >50 | 11 | -0.714 | -0.983 | -0.444 | <0.001 | 80.98% | <0.001 | -0.342 | -0.85 | 0.169 | 0.18 |
|  | Publication month | Feb/Mar | 9 | -0.452 | -0.712 | -0.192 | <0.001 | 66.65% | 0.002 | Reference |  |  |  |
|  |  | April | 7 | -0.979 | -1.354 | -0.604 | <0.001 | 70.53% | 0.002 | -0.572 | -0.97 | -0.17 | 0.006 |
| **Procalcitonin** | Overall |  | 10 | 0.868 | 0.508 | 1.228 | <0.001 | 88.16% | <0.001 |  |  |  |  |
|  | Origin of patients | Others | 5 | 1.038 | 0.370 | 1.706 | <0.001 | 86.16% | <0.001 | *Reference* |  |  |  |
|  |  | Wuhan | 5 | 0.686 | 0.331 | 1.041 | <0.001 | 75.38% | 0.003 | -0.318 | -0.97 | 0.33 | 0.34 |
|  | Sample size | ≤50 | 3 | 0.768 | 0.334 | 1.203 | <0.001 | 0% | 0.80 | *Reference* |  |  |  |
|  |  | >50 | 7 | 0.903 | 0.459 | 1.348 | <0.001 | 88.62% | <0.001 | 0.054 | -0.72 | 0.83 | 0.89 |
|  | Publication month | Feb/Mar | 6 | 0.956 | 0.404 | 1.509 | <0.001 | 91.51% | <0.001 | *Reference* |  |  |  |
|  |  | April | 4 | 0.757 | 0.409 | 1.105 | <0.001 | 41.54% | 0.16 | -0.096 | -0.80 | 0.61 | 0.78 |
| **C-reactive protein** | Overall |  | 13 | 1.027 | 0.65 | 1.40 | <0.001 | 88.2% | <0.001 |  |  |  |  |
|  | Origin of patients | Others | 8 | 1.24 | 0.65 | 1.83 | <0.001 | 87.8% | <0.001 | *Reference* |  |  |  |
|  |  | Wuhan | 5 | 0.389 | 0.30 | 1.07 | <0.001 | 80.7% | <0.001 | -0.58 | -1.27 | 0.10 | 0.09 |
|  | Sample size | ≤50 | 3 | 0.831 | 0.341 | 1.322 | <0.001 | 0% | 0.58 | *Reference* |  |  |  |
|  |  | >50 | 10 | 1.08 | 0.651 | 1.512 | <0.001 | 82.3% | <0.001 | 0.37 | -0.55 | 1.29 | 0.42 |
|  | Publication month | Feb/Mar | 8 | 1.014 | 0.502 | 1.525 | <0.001 | 88.23% | <0.001 | *Reference* |  |  |  |
|  |  | April | 5 | 1.07 | 0.548 | 1.600 | <0.001 | 75.1% | 0.003 | 0.13 | -0.59 | 0.86 | 0.71 |

SMD: Standardized mean difference, LL: lower limit, UL: upper limit, $I^2$ : the ratio of true heterogeneity to total observed variation.
Significant values indicate significance at $P < 0.05$ .

### Meta-regression analysis

Considering the number of the included studies with severity, ICU admission, and mortality data was rather small, we performed meta-regression analyses for only five parameters (mentioned above) in studies comparing mild and severe disease (**Table 4)**.

For WBCs, higher difference between mild and severe cohorts was noted in Wuhan studies than other population (coefficient = 0.31, 95%CI = 0.03, 0.58, *p* = 0.029). Moreover, articles with larger sample size exhibited a wider variation of neutrophilic count between severe and non-severe cases (coefficient = 0.60, 95%CI = 0.20, 1.01, *p* = 0.003). For the same marker, later studies published in April also showed higher difference compared to those published in February and March (coefficient = 0.31, 95%CI = 0.06, 0.55, *p* = 0.014). In contrast, more reduction of lymphocytes was observed in April articles than earlier ones (coefficient = -0.57, 95%CI = -0.97, -0.17, *p* = 0.006).

### Publication bias

Publication bias was performed to the same five parameters with study count ≥10 (**Fig. S1)**. Visual inspection of the funnel plots suggested symmetrical distribution for all laboratory parameters tested. The Egger test (*p* > 0.1) confirmed that there was no substantial evidence of publication bias; Egger’s regression *p* values were 0.44, 0.50, 0.68, 0.56, and 0.22 for WBC, neutrophil count, lymphocyte count, procalcitonin, and CRP, respectively.

### Decision tree and Receiver Operating Characteristic (ROC) curve

To identify predictors for severity, decision tree analysis was applied using multiple laboratory results. High performance of classification was found with the usage of a single parameter; neutrophilic count identified severe patients with 100% sensitivity and 81% specificity at a cut-off value of >3.74 identified by the specified decision tree model. Further analysis of the area under the curve of input data is shown in **Table 5**.

**Table 5.** Receiver Operating Characteristics results.

| Lab test | AUC | Threshold | Sensitivity | Specificity | P-value |
| --- | --- | --- | --- | --- | --- |
| WBC | 0.801 ± 0.09 | 5.47 | 85.7 | 85.7 | **0.007** |
| Neutrophil | 0.831 ± 0.09 | 3.74 | 78.5 | 100 | **0.003** |
| Lymphocyte | 0.867 ± 0.06 | 0.98 | 81.2 | 87.5 | **<0.001** |
| Platelets | 0.836 ± 0.11 | 177.6 | 71.4 | 71.4 | **0.035** |
| PT | 0.583 ± 0.17 | 12.9 | 50.0 | 83.3 | 0.63 |
| Procalcitonin | 0.845 ± 0.09 | 0.06 | 80.0 | 90.0 | **0.007** |
| D-dimer | 0.876 ± 0.08 | 0.48 | 88.9 | 77.8 | **0.007** |
| CRP | 0.875 ± 0.08 | 38.2 | 84.6 | 92.3 | **0.001** |
| IL-6 | 0.632 ± 1.6 | 22.9 | 71.4 | 71.4 | 0.40 |

AUC: area under the curve, WBC: white blood cells, PT: prothrombin time, CRP: C-reactive protein, IL-6: interleukin 6. Bold values indicate significance at $P < 0.05$ .

### Trial sequential analysis

As elaborated by the decision tree algorithm for the role of neutrophilic count on decision-making to discriminate between COVID-19 patients with a mild and severe presentation, TSA was employed on that particular laboratory parameter to test for the presence of sufficient studies from which results were drawn. The sample size of studies containing neutrophilic count information and classifying cohorts into mild and severe COVID-19 infection accounted for a total of 1,558 subjects. TSA illustrated crossing of the monitoring boundary by the cumulative Z-curve before reaching the required sample size, suggesting that the cumulative proof was acceptable, and no additional future studies are needed to authenticate the significances (**Fig 2)**.

**Fig 2.**
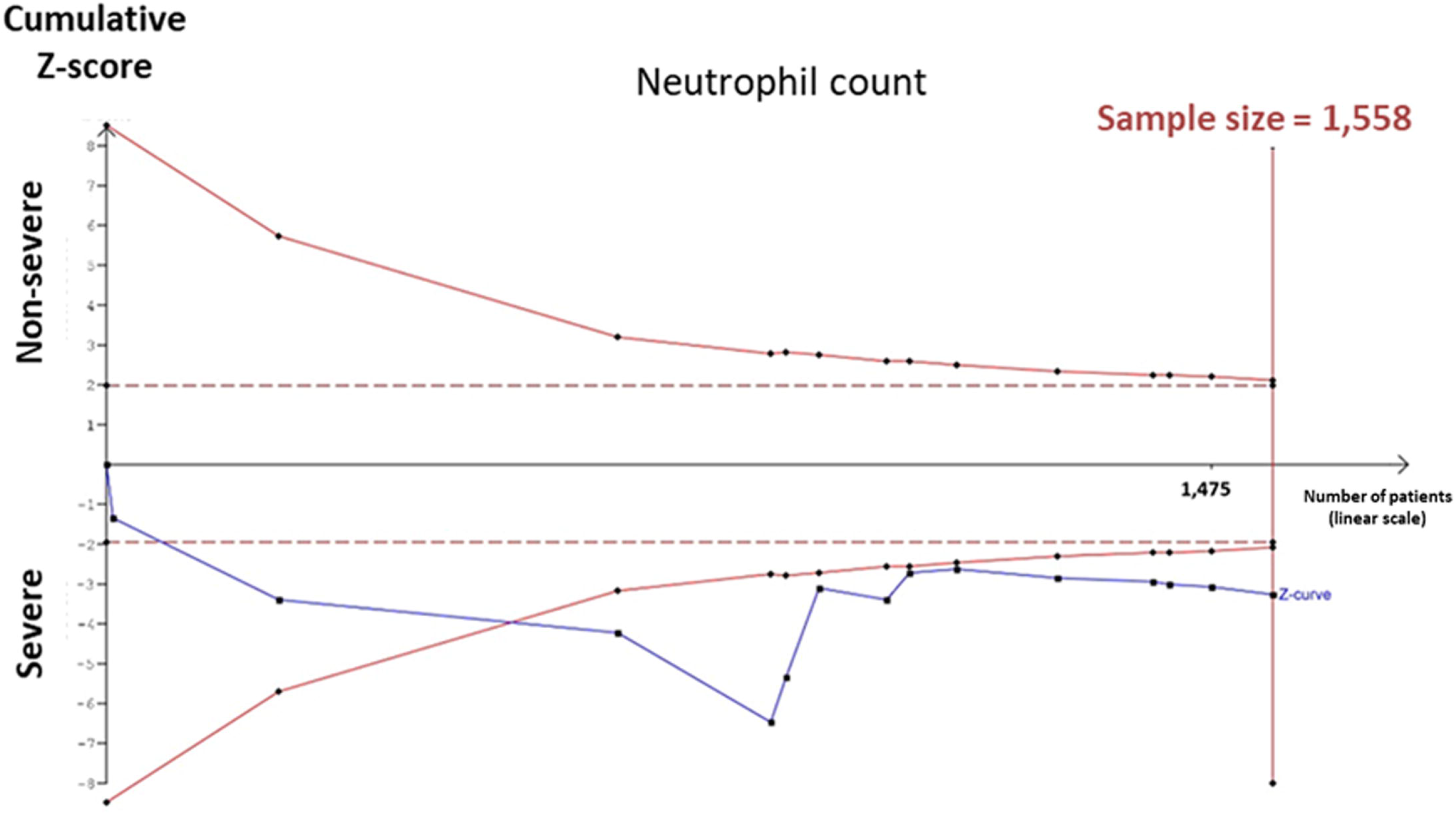
Trial sequential analysis. Trial sequential analysis (TSA) for the neutrophil count. The obtained sample size of the neutrophil count was 1558 subjects and the cumulative Z-curve crossed the monitoring boundary before reaching the required sample size, suggesting that the cumulative proof was reliable, and no additional trials are required to achieve the significances.

## Discussion

During the last few months, the prevalence of COVID-19 infection was increased daily among different countries overall in the world. Thus, the need to assess the disease severity and mortality are required to limit the pervasiveness of this pandemic [21]. A diverse of abnormal laboratory parameters including hematological, inflammatory as well as immunological markers thought to be raised throughout COVID-19 outbreak [2, 22]. In this comprehensive meta-analysis, our team attempted to interpret the distinct questions raised about the various spectrum of laboratory parameters associated with the severity and mortality of COVID-19. At the beginning of this workflow, our team investigated different hematological, inflammatory, and immunological variables of 6320 patients diagnosed with COVID-19. Our findings using random-effect models revealed increased levels of WBCs and neutrophil counts that were significantly associated with higher odds ratio among severe, ICU admission and Expired patients with COVID-19. On the contrary, the levels of lymphocyte and platelet counts were lowered among severe and expired patients with COVID-19. Also, we observed depletion in quantities of CD4^+^ T cells and CD8^+^ T cells among severe and mortality patients.

Nevertheless, in patients with the COVID-19 outbreak, the WBC count can vary [23]. Other reports indicated that leukopenia, leukocytosis, and lymphopenia have been reported, although lymphopenia appears most common [24, 25]. Another study supported that lymphopenia is an effective and reliable indicator of the severity and hospitalization in COVID-19 patients [26]. The additional report suggested that COVID-19 illness might be implicated with CD4^+^ and CD8^+^ T cells depletion through acting on lymphocytes, especially T lymphocytes [27]. A recent meta-analysis study discovered that the severity among COVID-19 patients might correlate with higher levels of WBCs count and lower levels of lymphocyte, CD4^+^ T cells, and CD8^+^ T cells counts [22]. In this respect, we could speculate that the depletion in the number of lymphocytes count is directly proportional with the severity of COVID-19 infection and the high survival rate of the disease is associated with the ability to renovate lymphocyte cells, particularly T lymphocytes which are crucial for destroying the infected viral particles [28]. During disease severity, remarkable thrombocytopenia was observed and confirmed by Lippi and his colleagues that revealed a reduction of platelet count among severe and died patients with COVID-19 supporting that thrombocytopenia could consider as an exacerbating indicator during the progression of the disease [29]. Therefore, our findings could support Shi et al. conclusion that high WBC count with lymphopenia could be considered as a differential diagnostic criterion for COVID-19 [30].

Considering coagulation profile, our team observed a prolonged in most coagulation markers among severe, ICU and expired patients, especially prothrombin time, fibrinogen, D-dimer, but with normal proportions of activated partial thromboplastin time (APTT) that could focus the light on the pathogenesis of COVID-19 infection through interfering with extrinsic coagulation pathway. A recently published report concluded similar findings in the form of observation of higher levels prothrombin time, D-dimer along fibrin degradation products among non-survival compared with survival patients [31].

Numerous studies illustrated the pathogenesis action of COVID-19 with the induction of cytokine storm throughout the progressive phase of the infection [22, 32, 33]. The generation of cytokine storm within COVID-19 patients required increased levels of IFN-γ and IL-1β that could stimulate the cellular response of T helper type 1 (Th1) which has a crucial function in the acceleration of specific immunity against COVID-19 outbreak [32]. Due to the elevated levels of IL-2R and IL-6 accompanied by the advancement of COVID-19, several cytokines secreted by T helper type 2 (Th2) cells that could neutralize the inflammatory responses including IL-4 and IL-10 [22, 32]. Our findings revealed a significantly associated with elevated levels of anti-inflammatory cytokines involving IL-6 and IL-10 among severe and expired patients with COVID-19. A recent study indicated a similar assumption with these findings and identified elevated levels of IL-6 and IL-10 among non-survived compared with survived patients [9]. Another confirmation of this conclusion is confirmed by a newly published meta-analysis report that indicated an exaggerated elevation of IL-6 and IL-10 throughout the severe level of COVID-19 infection [22].

Concerning the inflammatory markers associated with the COVID-19 pandemic, this comprehensive meta-analysis study observed higher concentrations of C-reactive protein (CRP) and procalcitonin besides elevated erythrocyte sedimentation rate (ESR) levels among severe and expired patients with COVID-19. Recently, Henry et al. established a meta-analysis survey and corroborated this finding with a higher significance of CRP and procalcitonin levels [22]. Other recent reports identified higher levels of CRP among severe patients with COVID-19 infection [26]. An additional meta-analysis survey established based on four recent articles indicated prolonged levels of procalcitonin among severe patients with COVID-19 [34]. In this respect, we might speculate the potential role of procalcitonin as a prognostic biomarker during the severe status of COVID-19. Finally, our team revealed increased levels of serum ferritin among non-survived patients compared with survived patients, and this significant outcome was observed in another meta-analysis study among severe and non-survival patients with COVID-19 infection [22].

This comprehensive meta-analysis confronted several limitations that raised throughout the processing of the outcomes. First, the insufficient laboratory data concerning the interest of design causing the increasing bias among different covariates. Second, the variation in the characteristics among different articles concerning the severity and survival of COVID-19. Third, the small sample sizes of some studies besides most of the concerned articles were established within China, especially Wuhan. Finally, there was an observed publication bias and heterogeneity in this comprehensive meta-analysis.

## Conclusion

In conclusion, several laboratory parameters could associate with the severity and mortality of COVID-19 infection and should be screened and measured continuously during the progression of this pandemic. These parameters included WBCs count, lymphocytes, platelet count, prothrombin time, D-dimer, and fibrinogen. Also, various interleukins could serve as anti-inflammatory markers such as IL-6, and IL-10 and should be evaluated. The estimation of other inflammatory biomarkers like CRP and procalcitonin could be helpful in the monitor the severity of the disease.

## Data Availability

All data are available in the manuscript and the supplementary materials.

## Acknowledgments

We thank all authors who provided published information for our meta-analysis.

## Supporting Materials

**- Table S1 PRISMA Checklist**.

**- Fig S1 Publication bias**

Funnel plot of standard error by the standardized difference in means for (A) White blood cells, (B) Neutrophil count, (C) Lymphocyte count, (D) Procalcitonin, and (E) C-reactive protein. The standard error provides a measure of the precision of the effect size as an estimate of the population parameter. It starts with zero at the top. Studies with smaller sample sizes produce less precise estimated effects with a broader base. The pooled estimated effects would be expected to scatter symmetrically around the total overall estimate of the meta-analysis (represented by the vertical line). Each circle represents a study (black circle). In the case of asymmetry, Duval and Tweedie’s trim and fill method predict the missing studies (red circle). Begg’s and Egger’s tests were performed. *P* values <0.1 were set to have a significant bias.

